# Mediation analysis in longitudinal data: an unbiased estimator for cumulative indirect effect

**DOI:** 10.64898/2026.04.18.26351189

**Authors:** Yi Li, Howard Cabral, Yorghos Tripodis, Jiantao Ma, Daniel Levy, Roby Joehanes, Chunyu Liu, Jungwun Lee

## Abstract

Mediation analysis quantifies how an exposure affects an outcome through an intermediate variable. We extend mediation analysis to capture the cumulative effects of longitudinal predictors on longitudinal outcomes. Our proposed model examines how mediators transmit the effects of the current and previous exposure on the current outcome. We construct a least-squared estimator for cumulative indirect effect (CIE) and used three approaches (exact form, delta method, and bootstrap procedure) to estimate its standard error (SE). The estimator of CIE is unbiased with no unmeasured confounding and independent model errors between mediator model and outcome model at all time points, as shown in statistical inference and in simulations. While three SE estimates are numerically similar, bootstrap procedure is recommended due to its simplicity in implementation. We apply this method to Framingham Heart Study offspring cohort to assess if DNA methylation mediates the association of alcohol consumption with systolic blood pressure over two time points. We identify two CpGs (cg05130679 and cg05465916) as mediators and construct a composite DNA methylation score from 11 CpGs, which mediates for 39% of the cumulative effect. In conclusion, we propose an unbiased estimator for CIE. Future studies will investigate the missingness in mediators and outcomes.

## 1. Introduction

Mediation analysis has become increasingly important for examining pathways between predictors and outcomes through intermediate variables, offering deeper insights into the mechanisms underlying disease etiology. Since Baron and Kenny first introduced the concept of mediation analysis using simple linear regression in 1986^1^, the field has significantly evolved. In recent years, there has been increasing interest in mediation analysis with longitudinal data. Many studies^2, 3, 4, 5, 6, 7^ used structural equation models for longitudinal mediation analysis, and other studies^8, 9^ used mixed models for repeated measures. The former allows complex relationships among exposures, mediators and outcomes, while the latter account for random effects. For example, latent variables are used in the structural equation model to capture the change of mediators and outcomes^10^, and linear mixed models have been applied to estimate natural direct and indirect effects conditional on random effect^8, 9^. However, relatively few studies have explored the cumulative effects of past and current exposures on the current outcome. Additionally, there is limited research on quantifying the cumulative indirect effect (CIE) through current mediators while accounting for historical exposures and outcomes. Addressing these gaps is important for understanding the long-term impact of exposures on outcomes.

Various methods^1, 11, 12, 13^ have been proposed for testing the significance of indirect effects. The most widely adopted approach involves dividing the estimated indirect effect by its standard error to obtain a z statistic, which is then compared to critical values from the standard normal distribution. Different methods have been developed to calculate standard error of indirect effect, such as difference-in-coefficients^14^, first-order solution^1^, second-order solution^11, 13^, and distribution of product^15^. While several studies have assessed different variance estimation methods by examining type I error rates and statistical power ^15, 16^, relatively few have evaluated the coverage probability of confidence intervals for indirect effects.

Therefore, our study aims to propose an unbiased estimator for CIE within the framework of longitudinal mediation analysis. CIE reflects the accumulative indirect effect of the exposure on the last outcome through mediators across all time points. We show that the proposed estimator is unbiased if the errors of outcome model and mediator model are independent, and unmeasured confounding. To our knowledge, this study is the first to specifically target the CIE, whereas other studies estimate indirect effect by averaging indirect effect over repeated measures, or time-lagged indirect effect. Our method extends the mediation framework developed by Baron and Kenny; thus, the CIE we estimate reflects an association rather than a causal effect. Furthermore, we estimate the variance of CIE by three approaches (exact variance method, delta method, and bootstrap procedure) and calculate the 95% confidence interval of CIE. The empirical coverage probability of the 95% confidence interval being close to 95% confirms that our variance estimate is robust and valid. Finally, we apply our proposed method to the Framingham Heart Study (FHS) offspring cohort to assess whether DNA methylation mediates the longitudinal association between alcohol consumption and blood pressure.

## 2. Methods

### 2.1 Definitions of mediation analysis

Mediation analysis is used to quantify the mechanisms through which an exposure or treatment (X) affects an outcome (Y) indirectly via one or more intermediate variables known as mediators (M). The total effect of X on Y can be decomposed into direct and indirect components. The direct effect refers to the portion of the exposure’s effect on the outcome that is not transmitted through any mediators, while the indirect effect represents the portion of the effect of X on Y that occurs through the mediator. This decomposition assesses whether and how the mediator alters the exposure-outcome relationship. In the mediation definition by Baron and Kenny, variable X, Y and M have the following relationship (**Figure 1. A**):

**Figure 1.**
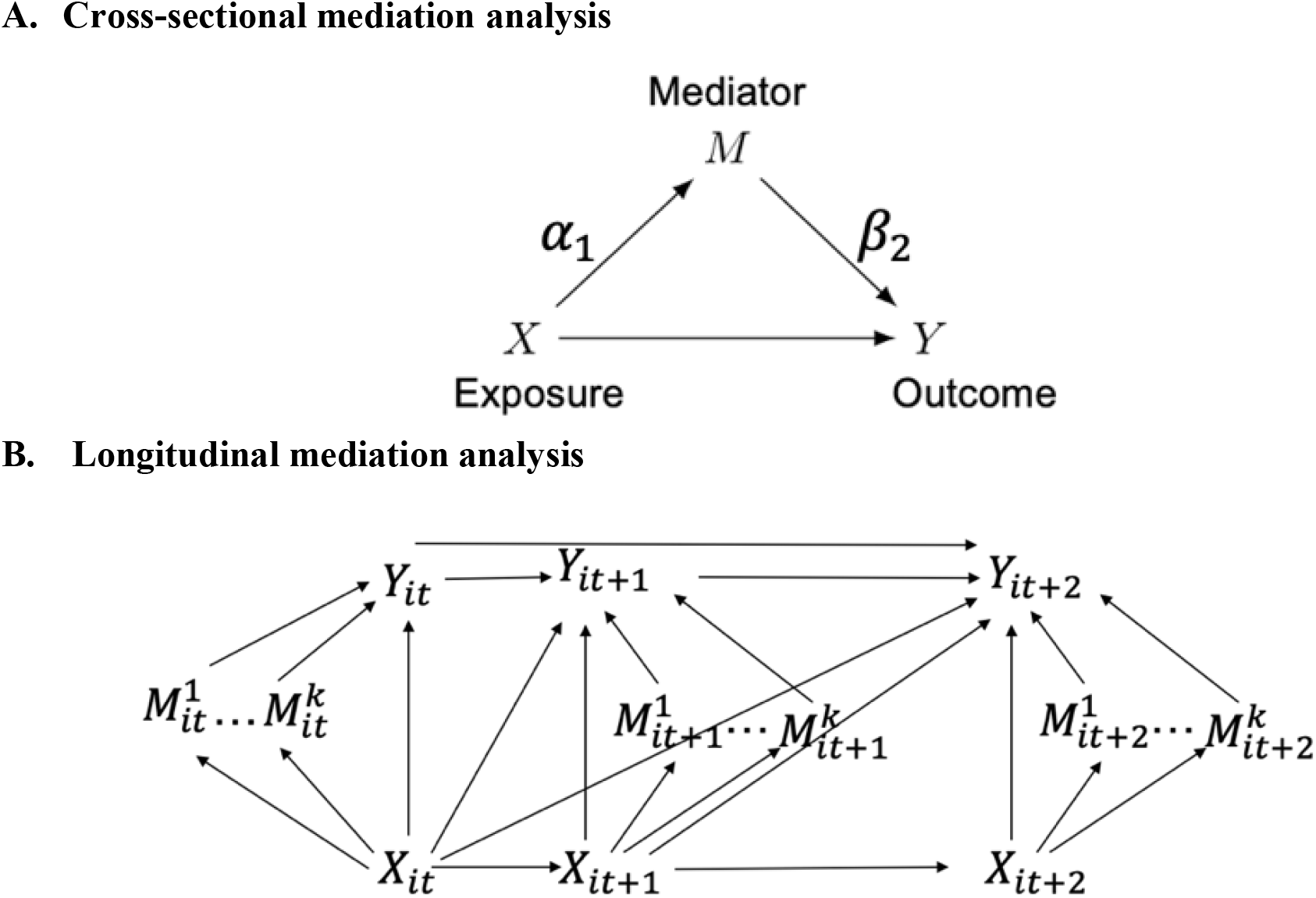
Directed acyclic graph of mediation analysis. **A. Cross-sectional mediation analysis** Definition of mediation analysis by Baron and Kenny: *α*_1_ is the association between exposure and mediator, *M* = *α*_0_ + *α*_1_*X* + *ε*_2_; *β*_2_ is the association between mediator and outcome, adjusting for exposure, *Y* = *β*_0_ + *β*_1_*X* + *β*_2_*M* + *ε*_3_. Indirect effect is *α*_1_*β*_2_. **B. Longitudinal mediation analysis** Outcome *Y*_*it*_ is affected by exposure *X*_*it*_ and multiple mediators 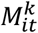. Outcome *Y*_*it*+1_ is affected by exposure *X*_*it*+1_, multiple mediators 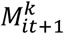, exposure *X*_*it*_ and outcome *Y*_*it*_. Outcome *Y*_*it*+2_ is affected by exposure *X*_*it*+2_ and multiple mediators 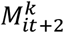, exposure *X*_*it*+1_, *X*_*it*_ and outcome *Y*_*it*+1_, *Y*_*it*_.

Total effect model: *Y* = *θ*_0_ + *θ*_1_*X* + *ε*_1_

Mediator model: *M* = *α*_0_ + *α*_1_*X* + *ε*_2_

Outcome model: *Y* = *β*_0_ + *β*_1_*X* + *β*_2_*M* + *ε*_3_ where [*θ*_0_, *α*_0_, *β*_0_] are intercept terms, and [*ε*_1_, *ε*_2_, *ε*_3_] are error terms. M represents a mediator if the following conditions are satisfied: (1) X is significantly associated with Y (i.e., *θ*_1_ ≠ 0); (2) X is significantly associated with M (i.e., *α*_1_ ≠ 0); (3) M is significantly associated with Y controlling for X (i.e., *β*_2_ ≠ 0). The total effect of X on Y in this situation is defined as *θ*_1_. The direct effect of X on Y is *β*_1_, and indirect effect is *α*_1_*β*_2_. This approach of calculating indirect effect is known as the product of method. Another approach is *θ*_1_ − *β*_1_, which is different method. Strong assumptions for mediation analysis include: (i) residuals in the mediator model *ε*_2_ and in the outcome are independent *ε*_3_, M and *ε*_3_ are independent; (ii) no X-M interaction in the outcome model; (iii) correct causal order (e.g., Y does not cause M) ; (iv) no misspecification due to unmeasured variables; (v) correct model specification.^1, 17^

Here we review three commonly used methods for obtaining the variance for the indirect effect, *α*_1_*β*_2_: delta method, exact variance method and bootstrap procedure. Sobel used multivariate delta method solution to calculate the variance 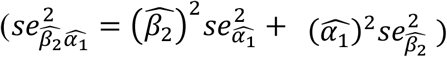. ^18^ Goodman^11^ and Aroian^13^ derived exact variance 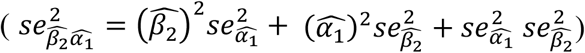 based on second order Taylor series. While these analytic approaches are widely used, they require normality of the outcome. Alternatively, bootstrap procedures can be used to obtain variance and confidence interval (CI) without distributional assumptions, at the cost of additional computation. In this work, we extend these ideas to CIE in a longitudinal setting and compare the performance of exact variance, delta-method, and bootstrap.

### 2.2 Unbiased estimator for cumulative indirect effect

A longitudinal mediation analysis with three time points (*t, t* + 1, *t* + 2) is shown in Figure 2.B as an example. Let *i* denotes subjects:1,2, …, *n*. Continuous exposure *X*, continuous outcome *Y*, and multivariate continuous mediators *M*^*k*^ are longitudinal variables. In addition, we assume the mediators measured at the same time point are independent (in other words, 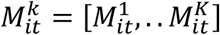 are mutually independent). Outcome *Y*_*it*_ is affected by exposure *X*_*it*_ and multivariate mediators 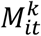. Outcome *Y*_*it*+1_ is affected by exposure *X*_*it*+1_, *X*_*it*_, multivariate mediators 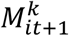 and previous outcome *Y*_*it*_. Outcome *Y*_*it*+2_ is affected by exposure *X*_*it*+2_, *X*_*it*+1_, *X*_*it*_, multivariate mediators 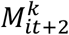 and previous outcome *Y*_*it*+1_, *Y*_*it*_. Note that we do not assume that (a) the mediator at different time points 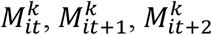 are independent from each other (that is,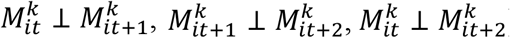), and (b) mediator and outcome not in the same time point are independent from each other (that is, 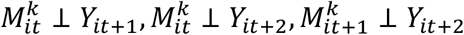).

**Figure 2.**
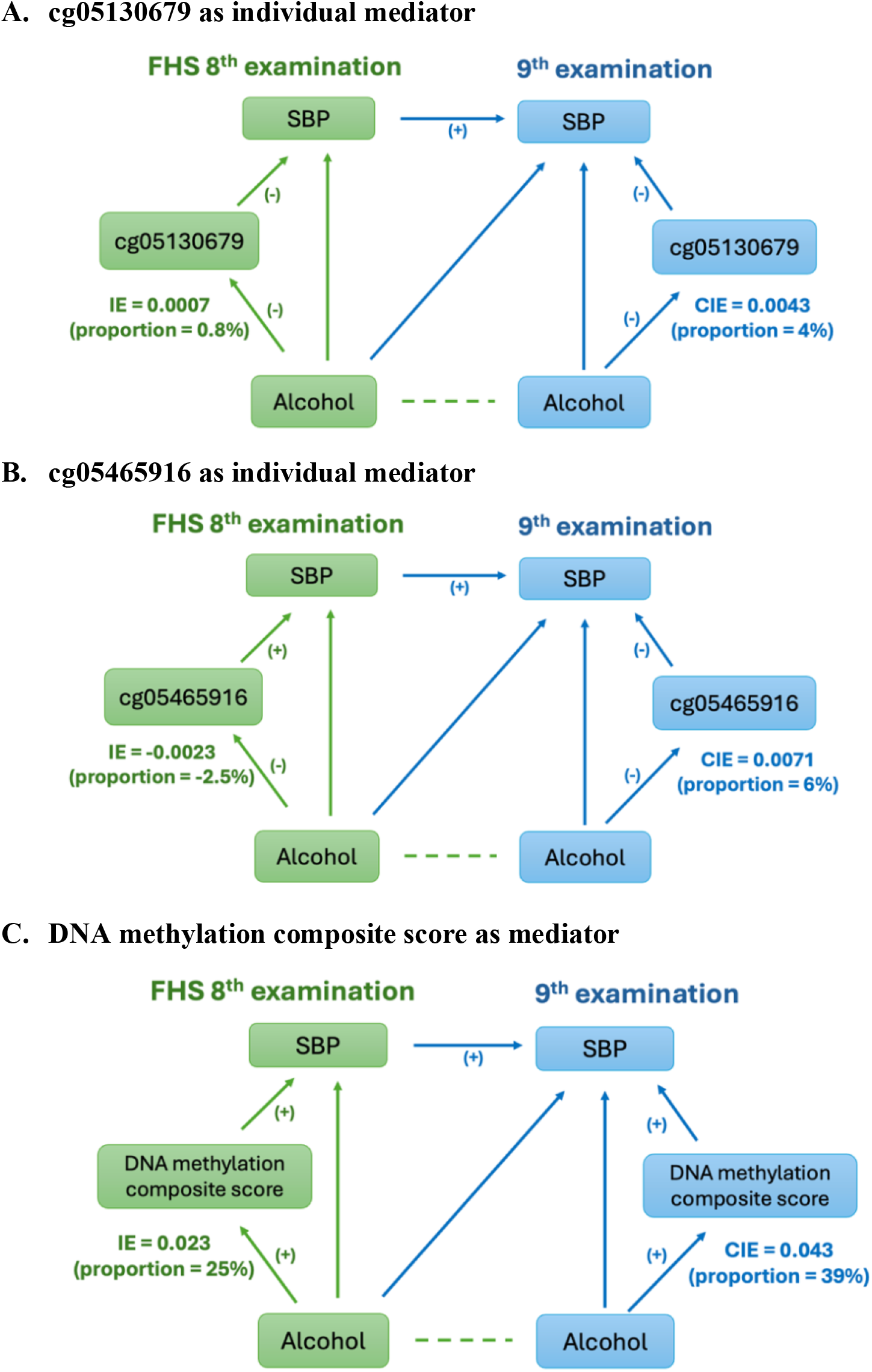
Mediation role of DNA methylation for the effect of alcohol consumption on systolic blood pressure. Mediation analysis was conducted in FHS offspring cohort at the 8^th^ and 9^th^ examinations (participant N=1,131). SBP: systolic blood pressure, IE: indirect effect at the 8^th^ examination; CIE: cumulative indirect effect across 8^th^ and 9^th^ examinations. Proportion: mediation proportion=cumulative indirect effect/total effect. (-), (+): direction of effect size for association between alcohol consumption and DNA methylation, DNA methylation and SBP, previous SBP and current SBP.

To analyze the relationships among the exposure, multiple mediators, and the outcome, we use the following modeling approach. First, we fit separate linear models for the exposure and each mediator. Second, we model all mediators on the outcome within one linear model.

Specially, the mediators are modeled as:

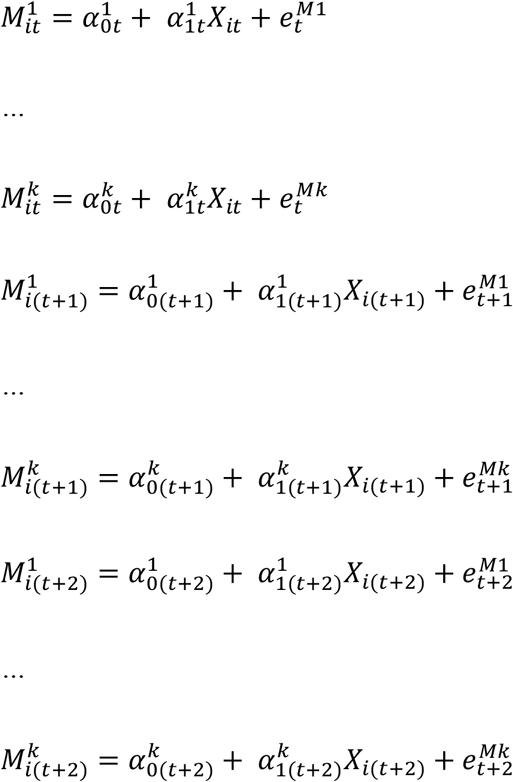

All mediators are then included jointly as predictors in the outcome model:

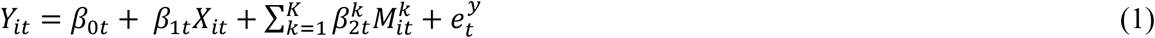

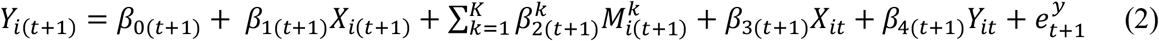

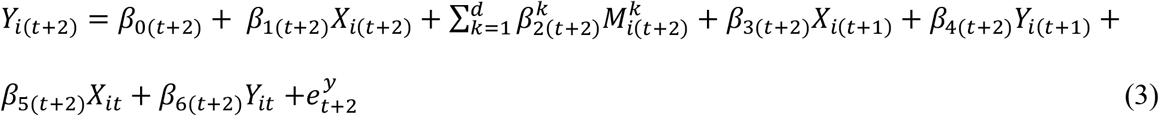

The direct effect of exposure *X*_*t*_ on outcome *Y*_*t*_ at time *t* is *β*_1*t*_ for per unit change in exposure *X*_*t*_. The specific indirect effect of exposure *X*_*t*_ on outcome *Y*_*t*_ through the k^th^ mediator 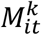 is 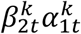. At time t+1, we substitute the model at time t (i.e., model (1)) into in model (2) to derive the cumulative effect of exposure *X*_*t*+1_, *X*_*t*_ and *Y*_*t*_ on outcome *Y*_*t*+1_, and details for derivation are in the Supplementary material S1. For per unit change in *X*_*t*+1_, *X*_*t*_ and *Y*_*t*_, the cumulative direct effect is *β*_1(*t*+1)_ + *β*_3(*t*+1)_ + *β*_4(*t*+1)_*β*_1*t*_ and CIE through k^th^ mediator 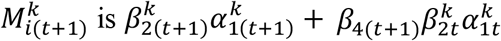. Similarly, we use model (1) and (2) to substitute it into model (3) to derive the cumulative effect of exposure *X*_*t*+2_ *X*_*t*+1_, *X*_*t*_, *Y*_*t*+1_ and *Y*_*t*_ on outcome *Y*_*t*+2_ . For per unit change in *X*_*t*+2_, *X*_*t*+1_, *X*_*t*_, *Y*_*t*+1_ and *Y*_*t*_, the cumulative direct effect is *β*_1(*t*+2)_ + *β*_3(*t*+2)_ + *β*_4(*t*+2)_*β*_1(*t*+1)_ + *β*_4(*t*+2)_*β*_3(*t*+1)_ + *β*_4(*t*+2)_*β*_4(*t*+1)_*β*_1*t*_ + *β*_5(*t*+2)_ + *β*_6(*t*+2)_*β*_1*t*_, and CIE through k^th^ mediator 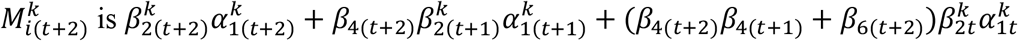. Using the law of total expectation, we show in Supplementary Section S2 that, the estimator for CIE is unbiased under independence of error terms in mediator and outcome models at all time points.

### 2.3 Variance of the cumulative indirect effect

We propose three different types of variance estimates for CIE: the delta method, exact variance form, and bootstrap procedure. Goodman developed the method to calculate the product of two independent random variables, and product of three independent random variables.^11^ Based on these formulas, we calculate exact variance at time t and t+1. Take one mediator as an example, we obtain the variance of the indirect effect at time t:

By delta method: 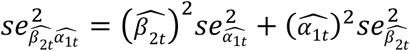

By exact variance form: 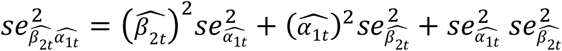

At time t+1, we obtain the variance:

By delta method: 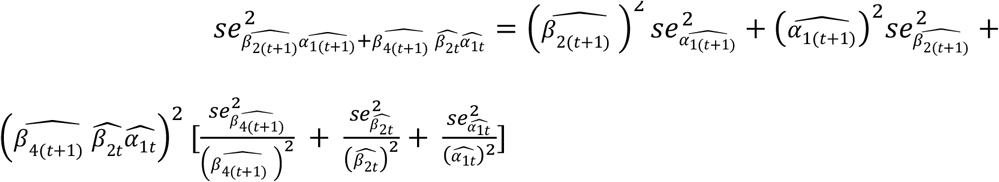

By exact method: 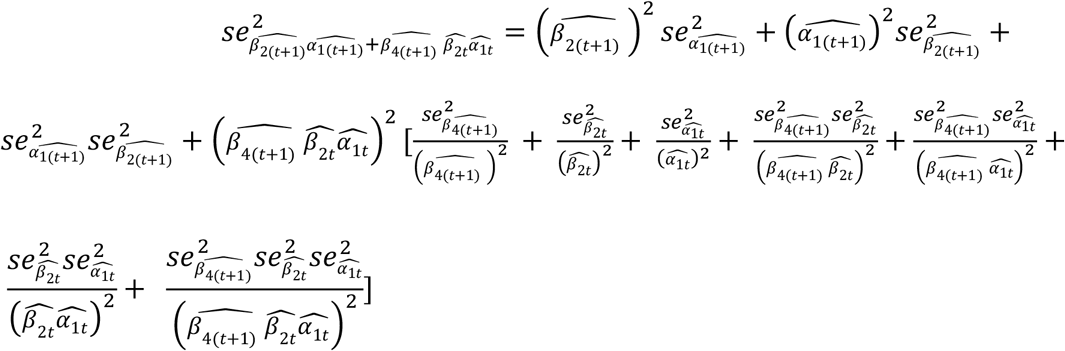

At time t+2, due to the complexity of the exact variance method, we obtain the variance by delta method: 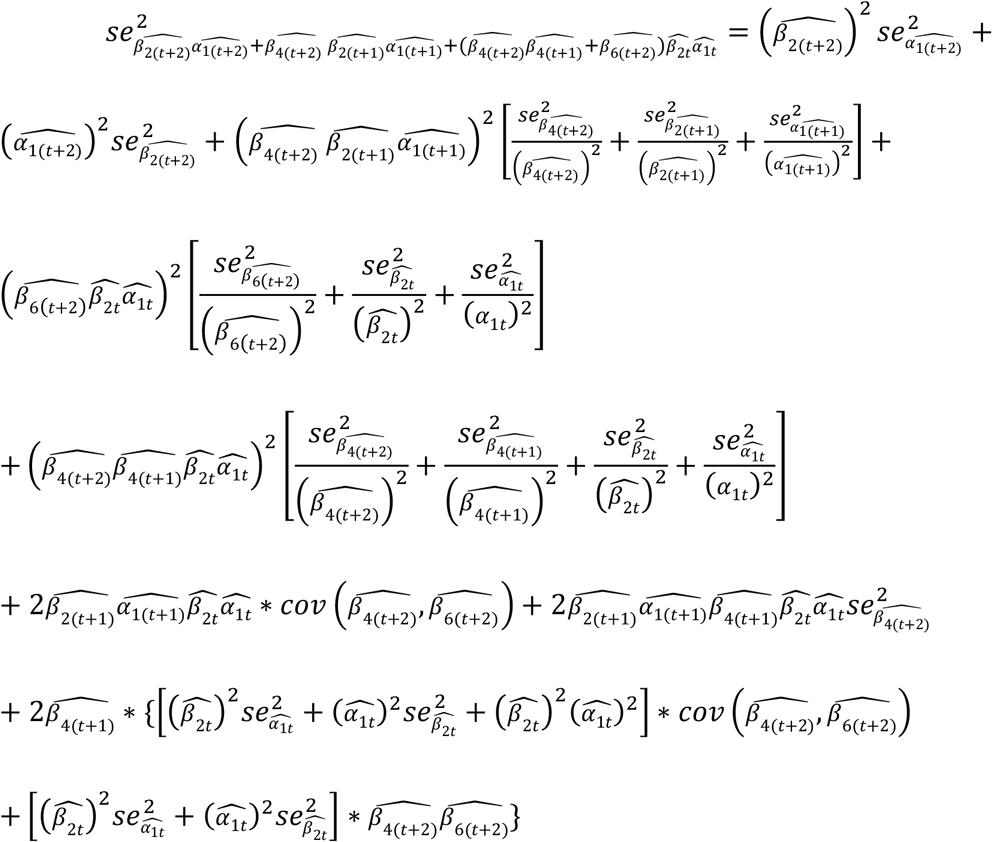

## 3. Simulation study design

In this section, we performed a simulation study to evaluate the estimator for the CIE and compared different estimators for variance of CIE.

### 3.1 Data generation

We generated data for 1000 participants at three time points. The exposure vector followed a multinormal distribution, (*X*_*t*_, *X*_*t*+1_, *X*_*t*+2_)^*T*^ ∼ *MNV*(***μ***, *∑*), with an auto-regressive correlation structure. The Pearson correlation between adjacent time points was set to either moderate or high correlated. The data-generating model specified a single mediator that was associated with both the exposure and outcome. At each time point, the variance explained by a current exposure in the mediator model was modest (*R*^2^ = 0.14). The variance explained by the exposure at all time points, current mediator, and the outcome at previous time points in the outcome model was either medium (*R*^2^ ≈ 0.56) or low (*R*^2^ ≈ 0.15).

We designed four scenarios according to Pearson correlation between the exposure at three time points and variance explained in the outcome model. Parameter values under each scenario were presented in Supplementary Material S3. The variance estimators of CIE by exact variance form and delta method are described in the Method section. In addition, we used bootstrap procedure to estimate CIE variance. We performed 1,000 independent simulation replicates to evaluate the estimator for CIE and its variance. Furthermore, we conducted a sensitivity analysis using a smaller sample size (n = 100) to compare with the primary analysis based on a larger sample size (n = 1000). Moreover, we compared our proposed cumulative modeling approach with the cross-lagged panel model in the simulated scenario 1 setting (**Supplementary Material S4**). Finally, we implemented a sensitivity analysis by including a confounder between the exposure and the mediator to investigate the effect of unmeasured confounders on the estimation of CIE (**Supplementary Material S5**).

### 3.2 Results of simulation study

We used absolute bias percent 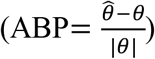 and root mean squared error 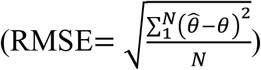 to evaluate the accuracy of the estimator for CIE. We also used coverage probability to compare 95% CI from different variance estimators.

#### Evaluation of point estimate

The bias of CIE estimates consistently remained below 1% (**Table 1**), and RMSE were low across all four scenarios and all three time points, varying from 0.014 to 0.025. These results clearly demonstrate that neither exposure correlations nor proportion of variance explained in the outcome model substantially influenced the accuracy of estimated CIE. These findings are consistent with the theoretical unbiasedness of the estimator under independent model errors in mediator and outcome models. The smaller sample size (n=100) showed relatively higher absolute bias percentages across scenarios and time points, ranging from -3.29% to 1.20%. Similarly, RMSE values (0.046 to 0.081) were larger when sample sizes were smaller. We conclude that the estimation of CIE was reasonably accurate and precise, even with the smaller sample size (**Table 1**).

**Table 1.** Bias and root mean square error for cumulative indirect effect.

| Sample size | Time | Absolute bias percent |  |  |  | RMSE |  |  |  |
| --- | --- | --- | --- | --- | --- | --- | --- | --- | --- |
|  |  | Scenario1 | Scenario2 | Scenario3 | Scenario4 | Scenario1 | Scenario2 | Scenario3 | Scenario4 |
| n=1000 | t | -0.42% | 0.21% | -0.60% | 0.16% | 0.022 | 0.023 | 0.014 | 0.014 |
|  | t+1 | -0.31% | 0.28% | 0.37% | -0.10% | 0.024 | 0.025 | 0.015 | 0.015 |
|  | t+2 | -0.14% | 0.16% | 0.73% | 0.65% | 0.025 | 0.025 | 0.015 | 0.015 |
| n=100 | t | -1.12% | 1.20% | -2.61% | 0.97% | 0.073 | 0.074 | 0.046 | 0.047 |
|  | t+1 | -0.27% | 0.18% | -1.74% | -0.98% | 0.078 | 0.078 | 0.047 | 0.047 |
|  | t+2 | -0.36% | 0.85% | -3.29% | -0.28% | 0.081 | 0.081 | 0.049 | 0.049 |

Scenario 1: moderate correlated exposure at three time points, large variance explained in the outcome model
Scenario 2: highly correlated exposure at three time points, large variance explained in the outcome model
Scenario 3: moderate correlated exposure at three time points, small variance explained in the outcome model
Scenario 4: highly correlated exposure at three time points, small variance explained in the outcome model
RMSE: mean square error

#### Comparison of variance estimation

Three estimators for the CIE variance (i.e., estimators estimated from the exact variance, delta method, and bootstrap procedure methods) yielded similar estimates. When the exposure at all time points, current mediator, and the outcome at previous time points explained a large proportion of variance in outcome model (scenario 1 and 2), the CIE variance effect was small: 0.0005 (time t), 0.0006 (t+1), and 0.0007 (t+2). Such a trend was similar when the proportion of variance from the outcome model was small (scenario 3 and 4).

While the exact variance and delta methods involve complex calculations and become cumbersome in longitudinal studies with multiple mediators, the bootstrap procedure is relatively simple to implement, requires fewer assumptions, and provides reliable estimates, making it well suited for analyzing complex data. As a result, we recommend using the bootstrap standard errors in the real data analysis.

Across all four scenarios, the three methods achieved coverage probabilities close to 0.95 at each time point. For the larger sample size (n=1000), coverage probabilities ranged between 0.93 and 0.97, indicating excellent accuracy. The smaller sample (n=100) showed slightly lower but still acceptable coverage (0.92 to 0.96) (**Table 2**). This finding demonstrated the reliability and robustness of our estimator in achieving appropriate confidence interval coverage. In the sensitivity analysis of unmeasured confounder, the bias and RMSE increased progressively, and coverage probability decreased as increasing confounding strength (**Supplementary Figure 2 and 3**). This finding suggested that the proposed CIE estimator lacked robustness in the presence of unmeasured confounders.

**Table 2.** Coverage probability of cumulative indirect effect.

|  | sample size | Time | Exact<br>variance | Delta<br>method | Bootstrap |
| --- | --- | --- | --- | --- | --- |
| Scenario1 | n=1000 | t | 0.96 | 0.96 | 0.95 |
|  |  | t+1 | 0.96 | 0.96 | 0.95 |
|  |  | t+2 | \ | 0.97 | 0.94 |
|  | n=100 | t | 0.94 | 0.94 | 0.94 |
|  |  | t+1 | 0.95 | 0.95 | 0.95 |
|  |  | t+2 | \ | 0.95 | 0.94 |
| Scenario2 | n=1000 | t | 0.95 | 0.95 | 0.95 |
|  |  | t+1 | 0.95 | 0.95 | 0.94 |
|  |  | t+2 | \ | 0.96 | 0.94 |
|  | n=100 | t | 0.94 | 0.94 | 0.94 |
|  |  | t+1 | 0.95 | 0.94 | 0.94 |
|  |  | t+2 | \ | 0.96 | 0.95 |
| Scenario3 | n=1000 | t | 0.93 | 0.93 | 0.93 |
|  |  | t+1 | 0.94 | 0.94 | 0.94 |
|  |  | t+2 | \ | 0.96 | 0.95 |
|  | n=100 | t | 0.93 | 0.92 | 0.93 |
|  |  | t+1 | 0.94 | 0.94 | 0.94 |
|  |  | t+2 | \ | 0.94 | 0.94 |
| Scenario4 | n=1000 | t | 0.95 | 0.95 | 0.95 |
|  |  | t+1 | 0.94 | 0.94 | 0.93 |
|  |  | t+2 | \ | 0.96 | 0.95 |
|  | n=100 | t | 0.94 | 0.93 | 0.92 |
|  |  | t+1 | 0.96 | 0.95 | 0.95 |
|  |  | t+2 | \ | 0.95 | 0.94 |

Scenario 1: moderate correlated exposure at three time points, large variance explained in the outcome model
Scenario 2: highly correlated exposure at three time points, large variance explained in the outcome model
Scenario 3: moderate correlated exposure at three time points, small variance explained in the outcome model
Scenario 4: highly correlated exposure at three time points, small variance explained in the outcome model

## 4. Real data application

In this section, we demonstrated the application of our model to explore whether DNA methylation mediates the cumulative effect of total alcohol consumption on the systolic blood pressure (SBP), using the Framingham Heart Study (FHS) data. We focused on alcohol-related DNA methylation at cytosine-phosphate-guanine (CpG) sites that were previously identified in a large epigenome-wide association study (EWAS).^19^

### 4.1 Data description

#### Study participant

The FHS offspring cohort began in 1971, with 5,124 participants undergoing in-person examinations every four to eight years.^20, 21^ These health assessments gathered comprehensive demographic information and clinical data, including risk factors for cardiovascular and neurodegenerative diseases. During each examination, a physician measured blood pressure twice in succession using a mercury sphygmomanometer following a standardized procedure. The average of these two readings was used as the final blood pressure value.^22^ This study used FHS offspring cohort data at the 8^th^ (2005-2008) and 9^th^ (2011-2014) examination cycles.

#### Alcohol consumption

Alcohol consumption data were gathered using questionnaires administered by FHS technicians.^23^ Participants reported the typical frequency of consuming three types of alcoholic beverages (i.e., beer, wine, and liquor). A standard drink was defined as one 12-ounce beer, one 4-ounce glass of wine (red or white), or one 1.5-ounce serving of 80-proof liquor, with each drink containing roughly 14 grams of ethanol. We calculated daily grams of each of alcoholic beverage type and then calculated their sum as the total alcohol consumption.

#### DNA methylation measurement

DNA was extracted from whole blood samples. DNA methylation at CpG sites was assessed using the Infinium BeadChip platform (Illumina, San Diego, CA, USA). During the 8^th^ examination, approximately 440,000 CpG sites measured from 2,427 participants were analyzed with the Human Methylation 450K array. At the 9^th^ examination, nearly 859,000 CpG sites were analyzed the Human Methylation EPIC (850K) array from 1,260 participants. Approximately 93% of the CpG sites included in the 450K array are also represented on the EPIC array.^24^ The quality control process were previously described.^19^ For each CpG site, the methylation level was expressed as a β value, representing the proportion of methylated signal intensity.

#### Alcohol consumption-associated CpG sites

Liu et al. (2018) conducted an EWAS across 13 cohorts (n=13,317). Meta-analysis identified 363 alcohol related CpG sites in European ancestry samples (n=9,643) and 165 in African ancestry samples (n=2,423).^19^ Among the ones identified in European ancestry samples, 332 CpG sites were measured in FHS. Totally, 1,131 participants had alcohol consumption, SBP and DNA methylation measured at both 8^th^ and 9^th^ examinations.

### 4.2 Model Specification

Numerous studies have shown higher alcohol consumption was associated with higher SBP.^25, 26, 27, 28^ Building on previously reported associations of identifying 332 CpG sites associated with alcohol consumption, we tested whether DNA methylation at the 9^th^ examination mediated the cumulative association between alcohol consumption over two exams and SBP at the 9^th^ examination. Alcohol consumption was the exposure, SBP was outcome, and the DNA methylation levels at selected CpG sites were mediators. Covariates included age, sex, body mass index, white blood cells, and technical covariates for DNA methylation measurement and smoking status. We modeled each CpG site individually:

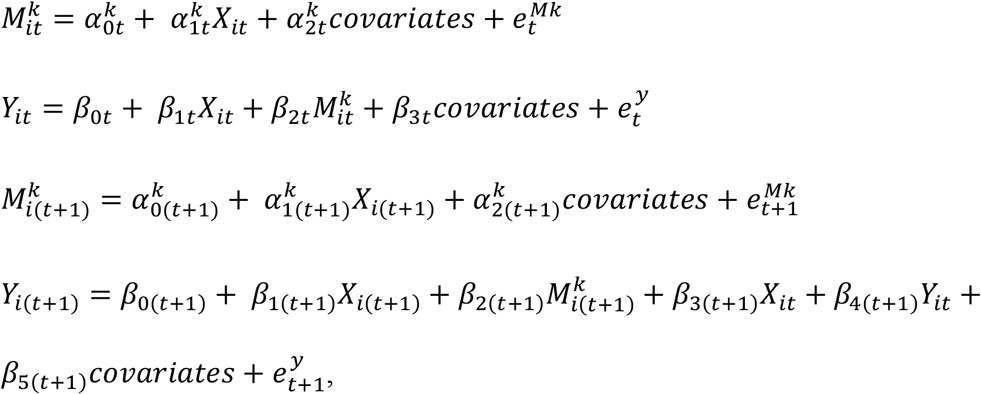

where t and t+1 were the FHS 8^th^ and 9^th^ examination, respectively; 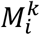 was the k^th^ CpG site. All the continuous variables were scaled with mean at zero and standard deviation (SD) at one for easier interpretability. We established the following criteria for selecting mediators. First, at the 9^th^ examination, the association of alcohol consumption with 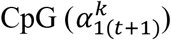 and the association of CpG with SBP (*β*_2(*t*+1)_) have identical direction (i.e., both positive or both negative). Second, CpG was significant associated with SBP (*β*_2(*t*+1)_ ≠ 0, at p<0.05) at the 9^th^ examination. At this stage, we did not adjust for multiple testing because the analysis is exploratory and restricted to CpGs previously associated with alcohol.

In addition, we constructed a composite DNA methylation score by applying elastic net regression (α = 0.5, λ = 2.2) to identify CpG sites associated with SBP. Here, α = 0.5 is a common choice in elastic net regression, and λ = 2.2 is the optimal value under α = 0.5. Due to the strong association between alcohol consumption and SBP and weak association between individual CpG and SBP, alcohol intake was excluded from the elastic net model to avoid penalizing and excluding relevant CpG sites. Based on selected CpG sites, we constructed a weighted score defined as 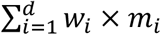, where d is total number of selected CpG site from elastic net regression, *m*_*i*_ is the raw value of DNA methylation at the i^th^ CpG site, and *w*_*i*_ is the corresponding coefficient from elastic net regression for this CpG site. This score was then standardized and utilized as a mediator in the mediation analysis, adjusting for the same covariates used in the individual CpG site analyses.

### 4.3. Results from real data analysis

With the criteria described in the model specification, we identified two CpG sites (cg05130679 and cg05465916) demonstrated marginal significant (P value < 0.10) CIE. At the 9^th^ examination, cg05130679 was negatively associated with alcohol drinking (beta = -0.07, *P* value = 0.02) and SBP (beta = -0.06, *P* value = 0.04). The CIE across two examinations was 0.0043 (*P* value = 0.08) with a mediation proportion of 4%. This suggested that 4% of cumulative effect of alcohol drinking across two examinations on the SBP was mediated by cg05130679. Notably, this CIE was substantially greater than indirect effect estimated from the 8^th^ examination alone (indirect effect = 0.0007, *P* value = 0.7) (**Figure 2. A**). Similarly, at the 9^th^ examinations, cg05465916 was both negatively associated with alcohol drinking (beta = -0.13, *P* value = 3E-6) and SBP (beta = -0.06, *P* value = 0.04). The CIE of cg05465916 was 0.0071 (*P* value = 0.09), and the mediation proportion was 6%. In contrast, at the 8^th^ examination, the indirect effect of cg05465916 was smaller (indirect effect= -0.0023, *P* value= 0.4) (**Figure 2. B**). Above results indicated that cumulative increased alcohol drinking was related to lower level at cg05130679 and cg05465916 site, and these decreased CpG levels were related to increased SBP.

Elastic net regression selected 11 CpG sites associated with SBP (**Supplementary Table 1**). At the 9^th^ examination, one SD higher level of alcohol consumption was associated with 0.32 SD rise in DNA methylation composite score (*P* value = 6×10^-42^), and one SD increase in DNA methylation composite score was associated with 0.1SD increase in SBP (*P* value = 0.006). The CIE across two examinations was 0.043 (*P* value = 0.001), indicating that DNA methylation composite score mediated the 39% of the cumulative effect of alcohol consumption on SBP. At the 8^th^ examination, DNA methylation composite score mediated for 25% effect (**Figure 2. C**). These findings suggested that the mediating role of these CpG sites may became more apparent when considering the cumulative impact of alcohol consumption on SBP over time rather than at a single time point.

## 5. Discussion

We proposed an unbiased estimator for CIE in longitudinal mediation analysis, capturing how repeated exposures influence a current outcome through current mediators and addressing a gap in existing methods that ignore cumulative effects. We compared several approaches for estimating the variance of the CIE and then applied the method to assess whether DNA methylation mediates the cumulative association between alcohol consumption and systolic blood pressure. Evidence from both individual CpG sites and a composite methylation score supports a role for DNA methylation in the alcohol–blood pressure relationship.

Our proposed method utilized longitudinal exposure, mediator, and outcome data to estimate the CIE of exposure on outcome through mediator at all time points. By incorporating more information over time, we were able to explore complex pathways underlying changes in these variables. To the best of our knowledge, this was the first study to specifically focus on the CIE in the longitudinal context. Other previous studies used mixed effect models to calculate average indirect effects over time^8, 9^, use structural equation models for indirect effect from time-lagged exposures and mediators, or used baseline exposure to predict longitudinal mediators and outcomes ^29, 30^. In contrast, our approach adjusted for exposure and outcome at all previous times and decomposes their effect into direct and indirect effect, thereby capturing the accumulating indirect effect. We proved that this estimator is unbiased under the assumption that the error terms in the outcome model and the mediator model at all time points are independent from each other. To estimate the CIE variance, we used three methods: exact variance approach, delta method, and bootstrap procedure. Bootstrap procedure was recommended due to its implement simplicity and no requirement for distribution. Subsequently, we calculated the coverage probability of 95% CI for the CIE. The high coverage probability (0.95) from all three methods indicated our variance estimator is valid.

Our method extends traditional mediation analysis, which typically formulated in terms of associations rather than causal relationships.^31, 32^ Causal mediation analysis, as developed by Robins and Greenland^31^ and Pearl^32^, defines natural direct and indirect effect using counterfactual framework and requires additional, strong assumptions. In this work, we focus on estimating cumulative indirect association within a linear-model framework and do not claim causal effects. Extending the CIE to a causal mediation setting would require careful handling of time varying confounding and cross-time dependencies, for example along the lines of the approach proposed by VanderWeele and Vansterrlandt^33^. Likewise, sensitivity analysis methods for unmeasured confounding in mediation analysis^34, 35^ could be adapted to assess the robustness of our findings.

We applied our proposed method to estimate the mediating effects of DNA methylation between alcohol consumption and SBP in the longitudinal setting in FHS. While individual CpGs might mediate only a small proportion of the total mediation effect, collectively they could account for a substantially larger proportion. Our findings show some consistency with prior studies. The negative association between alcohol consumption and most DNA methylation sites has been widely reported.^36, 37, 38^ In addition, prior studies^39, 40, 41^ have demonstrated negative association between DNA methylation and blood pressure. For example, Kaushik’s findings showed global DNA methylation was significantly negatively associated with both systolic and diastolic blood pressure.^39^ Similarly, in our study, ten out of eleven SBP-related CpG sites selected via elastic net regression showed negative associations. A cross-sectional study identified 66 CpG sites that significantly mediated the association between alcohol consumption and hypertension^42^. Two of these 66 CpG (cg06690548, cg00716257) was also detected in our study and used it for DNA methylation composite score. Six of 11 CpGs in our DNA methylation score and 14 of the 66 CpGs as individual mediators had moderately strong correlation (0.5∼0.67).

Our study has several limitations. First, our method is developed from traditional mediation analysis rather than causal mediation analysis. As discussed above, more work is needed to extend our method to causal mediation framework. Second, missing data remain a challenge in longitudinal mediation analysis, especially in the observational study; future analyses should incorporate improved imputation methods to address this issue. Third, although alcohol consumption and SBP were measured across multiple examinations in the FHS offspring cohort, DNA methylation data were only available at two time points. Additional repeated methylation measures would allow a more detailed assessment of its mediating role over time. Lastly, External validation of our findings in other independent cohorts would strengthen the generalizability and robustness of the mediation role of DNA methylation.

To sum up, our proposed method has addressed the gap of cumulative effects in longitudinal mediation analysis. The proposed estimator provides unbiased estimates for the cumulative indirect effect, capturing a more comprehensive relationship between exposures and outcomes over time. Additionally, we compared different estimation methods for variance. This new longitudinal mediation method deepens our understanding of the biological pathways underlying complex diseases.

## Data Availability

All data produced in the present work are contained in the manuscript.

## Abbreviations

ABP: Absolute bias percent
CI: Confidence interval
CpG: Cytosine-phosphate-guanine
FHS: Framingham Heart Study
RMSE: Root mean squared error
SBP: Systolic blood pressure
SE: Standard error
SD: Standard deviation

