## Supplementary_material for "Mediation analysis in longitudinal data: an unbiased estimator for cumulative indirect effect"

**S1. Derive cumulative indirect effect**

**S2. Proof of an unbiased estimator for the indirect effect**

**S3. Parameters for each simulation scenario**

**S4. Comparison to cross-lagged panel model**

**S5. Unmeasured confounding**

**Supplementary Table 1. Selected CpG and its coefficient from elastic net regression**

**Supplementary Table 2. Pearson correlation between exposures and unmeasured confounder**

**Supplementary Figure 1. Unmeasured confounders in the cumulative mediation analysis**

**Supplementary Figure 2. Bias for cumulative indirect effect under unmeasured confounder between exposure and mediator**

**Supplementary Figure 3. Coverage probability for cumulative indirect effect 95% confidence interval under unmeasured confounder between exposure and mediator**

### S1. Derive cumulative indirect effect

For the association between exposure and multiple mediators at a single time point, we model their relationship one by one. For the association between multiple mediators and outcome, we analyze them in one model. At time t, we can write out:

$$M_{it}^{1} = \alpha_{0t}^{1} + \ \alpha_{1t}^{1} X_{it} + e_{t}^{M1}$$

...

$$M_{it}^{k} = \alpha_{0t}^{k} + \ \alpha_{1t}^{k} X_{it} + e_{t}^{Mk}$$

$$Y_{it} = \beta_{0t} + \ \beta_{1t} X_{it} + \sum_{k=1}^{K} \beta_{2t}^{k} M_{it}^{k} + e_{t}^{y} = \beta_{0t} + \ \beta_{1t} X_{it} + \sum_{k=1}^{K} \beta_{2t}^{k} (\alpha_{0t}^{k} + \ \alpha_{1t}^{k} X_{it}) + e_{t}^{y}$$

$$= \beta_{0t} + \sum_{k=1}^{K} \beta_{2t}^{k} \alpha_{0t}^{k} + \ \beta_{1t} X_{it} + \sum_{k=1}^{K} \beta_{2t}^{k} \alpha_{1t}^{k} X_{it} + e_{t}^{y}$$

Thus, direct effect of exposure $X_t$ on outcome $Y_t$ at time $t$ is $\beta_{1t}$ for per unit change in exposure $X_t$. The specific indirect effect of exposure $X_t$ on outcome $Y_t$ through the k[th] mediator $M_{it}^{k}$ is $\beta_{2t}^{k}\alpha_{1t}^{k}$. The global indirect effect of exposure $X_t$ on outcome $X_t$ through all mediators is $\sum_{k=1}^{K} \beta_{2t}^{k}\alpha_{1t}^{k}$.

At time t+1, we can write:

$$M_{i(t+1)}^{1} = \alpha_{0(t+1)}^{1} + \ \alpha_{1(t+1)}^{1} X_{i(t+1)} + e_{t+1}^{M1}$$

...

$$M_{i(t+1)}^{k} = \alpha_{0(t+1)}^{k} + \ \alpha_{1(t+1)}^{k} X_{i(t+1)} + e_{t+1}^{Mk}$$

$$Y_{i(t+1)} = \beta_{0(t+1)} + \beta_{1(t+1)}X_{i(t+1)} + \sum_{k=1}^{K} \beta_{2(t+1)}^{k} M_{i(t+1)}^{k} + \beta_{3(t+1)}X_{it} + \beta_{4(t+1)}Y_{it} + e_{t+1}^{y}$$

$$= \beta_{0(t+1)} + \beta_{1(t+1)}X_{i(t+1)} + \sum_{k=1}^{K} \beta_{2(t+1)}^{k}(\alpha_{0(t+1)}^{k} + \alpha_{1(t+1)}^{k}X_{i(t+1)}) + \beta_{3(t+1)}X_{it} + \beta_{4(t+1)}(\beta_{0t}$$

$$+ \sum_{k=1}^{K} \beta_{2t}^{k}\alpha_{0t}^{k} + \beta_{1t}X_{it} + \sum_{k=1}^{K} \beta_{2t}^{k}\alpha_{1t}^{k}X_{it}) + e_{t+1}^{y}$$

The cumulative direct effect of exposure $X_{t+1}$, $X_t$ and $Y_t$ on outcome $Y_{t+1}$ is $\beta_{1(t+1)} + \beta_{3(t+1)} + \beta_{4(t+1)}\beta_{1t}$ for per unit change in $X_{t+1}$, $X_t$ and $Y_t$. The cumulative indirect effect of exposure $X_{t+1}$, $X_t$ and $Y_t$on outcome $Y_{t+1}$ through k[th] mediator $M_{i(t+1)}^{k}$ is $\beta_{2(t+1)}^{k}\alpha_{1(t+1)}^{k} + \beta_{4(t+1)}\beta_{2t}^{k}\alpha_{1t}^{k}$, the global cumulative indirect effect through all the mediators is $\sum_{k=1}^{K} (\beta_{2(t+1)}^{k}\alpha_{1(t+1)}^{k} + \beta_{4(t+1)}\beta_{2t}^{k}\alpha_{1t}^{k})$.

At time t+2, we can write:

$$M_{i(t+2)}^{1} = \alpha_{0(t+2)}^{1} + \alpha_{1(t+2)}^{1}X_{i(t+2)} + e_{t+2}^{M1}$$

...

$$M_{i(t+2)}^{k} = \alpha_{0(t+2)}^{k} + \alpha_{1(t+2)}^{k}X_{i(t+2)} + e_{t+2}^{Mk}$$

$$Y_{i(t+2)} = \beta_{0(t+2)} + \beta_{1(t+2)}X_{i(t+2)} + \textstyle\sum_{k=1}^{d} \beta_{2(t+2)}^{k}M_{i(t+2)}^{k} + \beta_{3(t+2)}X_{i(t+1)} + \beta_{4(t+2)}Y_{i(t+1)} + \beta_{5(t+2)}X_{it} + \beta_{6(t+2)}Y_{it} + e_{t+2}^{y}$$

$$=\beta_{0(t+2)} + \beta_{1(t+2)}X_{i(t+2)} + \textstyle\sum_{k=1}^{d} \beta_{2(t+2)}^{k}M_{i(t+2)}^{k} + \beta_{3(t+2)}X_{i(t+1)} + \beta_{4(t+2)}(\beta_{0(t+1)} + \beta_{1(t+1)}X_{i(t+1)} + \textstyle\sum_{k=1}^{K} \beta_{2(t+1)}^{k}\left(\alpha_{0(t+1)}^{k} + \alpha_{1(t+1)}^{k}X_{i(t+1)}\right) + \beta_{3(t+1)}X_{it} +$$

$\beta_{4(t+1)}(\beta_{0t} + \sum_{k=1}^{K} \beta_{2t}^{k}\alpha_{0t}^{k} + \ \beta_{1t}X_{it} + \sum_{k=1}^{K} \beta_{2t}^{k}\alpha_{1t}^{k}X_{it})) + \beta_{5(t+2)}X_{it} + \beta_{6(t+2)}(\beta_{0t} +$

$\sum_{k=1}^{K} \beta_{2t}^{k}\alpha_{0t}^{k} + \ \beta_{1t}X_{it} + \sum_{k=1}^{K} \beta_{2t}^{k}\alpha_{1t}^{k}X_{it}) + e_{t+2}^{y}$

Then we can get the cumulative direct effect of exposure $X_{t+2}$ $X_{t+1}$, $X_t$, $Y_{t+1}$ and $Y_t$ on outcome $Y_{t+2}$ is $\beta_{1(t+2)} + \beta_{3(t+2)} + \beta_{4(t+2)}\beta_{1(t+1)} + \beta_{4(t+2)}\beta_{3(t+1)} + \beta_{4(t+2)}\beta_{4(t+1)}\beta_{1t} + \beta_{5(t+2)} + \beta_{6(t+2)}\beta_{1t}$ for per unit change in $X_{t+2}$ , $X_{t+1}$, $X_t$ , $Y_{t+1}$ and $Y_t$ The cumulative indirect effect of exposure $X_{t+2}$, $X_{t+1}$, $X_t$, $Y_{t+1}$ and $Y_t$ on outcome $Y_{t+1}$ through k[th] mediator $M_{i(t+2)}^{k}$ is $\beta_{2(t+2)}^{k}\alpha_{1(t+2)}^{k} + \beta_{4(t+2)}\beta_{2(t+1)}^{k}\alpha_{1(t+1)}^{k} + (\beta_{4(t+2)}\beta_{4(t+1)} + \beta_{6(t+2)})\beta_{2t}^{k}\alpha_{1t}^{k}$, the global cumulative indirect effect through all the mediators is $\sum_{k=1}^{K}\{\beta_{2(t+2)}^{k}\alpha_{1(t+2)}^{k} + \beta_{4(t+2)}\beta_{2(t+1)}^{k}\alpha_{1(t+1)}^{k} + (\beta_{4(t+2)}\beta_{4(t+1)} + \beta_{6(t+2)})\beta_{2t}^{k}\alpha_{1t}^{k}\}$.

**S2. Proof of an unbiased estimator for the indirect effect**

We prove indirect effect estimator are unbiased. Since we assume mediators are independent from each other, both outcome model and mediator model are correctly specified. we here use one mediator as an example. At time t:

$M_{it}^{1} = \alpha_{0t}^{1} + \ \alpha_{1t}^{1}X_{it} + e_{t}^{M1}$,

$Y_{it} = \beta_{0t} + \ \beta_{1t}X_{it} + \beta_{2t}^{1}M_{it}^{1} + e_{t}^{y}$

From ordinary least squared method, $E(\widehat{\alpha_{1t}^{1}}|X = x) = \alpha_{1t}^{1}$, $E(\widehat{\beta_{2t}^{1}}|X = x, M = m) = \beta_{2t}^{1}$.

We assume that $e_{t}^{M1} \perp e_{t}^{y}$, then

$E(\widehat{\beta_{2t}^{1}}\widehat{\alpha_{1t}^{1}}) = E_{(\widehat{\alpha_{1}^{t}}, X,M)}(E_{\widehat{\beta_{2t}^{1}}}(\widehat{\beta_{2t}^{1}}\widehat{\alpha_{1t}^{1}}|X = x, M = m))$

$$= E_{\left(\widehat{\alpha_{1t}^1},\, X,M\right)}(\widehat{\alpha_{1t}^1} E_{\widehat{\beta_{2t}^1}}(\widehat{\beta_{2t}^1} | X = x, M = m))$$

$$= E_{\widehat{\alpha_{1t}^1}}(\widehat{\alpha_{1t}^1} \beta_{2t}^1)$$

$$= \beta_{2t}^1 E_{\widehat{\alpha_{1t}^1}}(\widehat{\alpha_{1t}^1}) = \beta_{2t}^1 E_X(E_{\widehat{\alpha_{1t}^1}}(\widehat{\alpha_{1t}^1} | X = x))$$

$$= \beta_{2t}^1\ \alpha_{1t}^1$$

Therefore $\widehat{\beta_{2t}^1}\widehat{\alpha_{1t}^1}$ is unbiased estimator for $\beta_{2t}^1\ \alpha_{1t}^1$.

Similarly, at time t+1,

$$M_{i(t+1)}^1 = \alpha_{0(t+1)}^1 + \ \alpha_{1(t+1)}^1 X_{i(t+1)} + e_{t+1}^{M1}$$

$$Y_{i(t+1)} = \beta_{0(t+1)} + \ \beta_{1(t+1)} X_{i(t+1)} + \sum_{k=1}^{K} \beta_{2(t+1)}^k M_{i(t+1)}^k + \beta_{3(t+1)} X_{it} + \beta_{4(t+1)} Y_{it} + e_{t+1}^y$$

Assuming $e_{t+1}^{M1} \perp e_{t+1}^y$, $e_t^y \perp e_{t+1}^y$, we can prove:

$$E\big(\widehat{\beta_{2(t+1)}^1 \alpha_{1(t+1)}^1} + \widehat{\beta_{4(t+1)}}\ \widehat{\beta_{2t}^1}\widehat{\alpha_{1t}^1}\big)$$

$$= E\big(\widehat{\beta_{2(t+1)}^1 \alpha_{1(t+1)}^1}\big) + E\big(\widehat{\beta_{4(t+1)}}\ \widehat{\beta_{2t}^1}\widehat{\alpha_{1t}^1}\big)$$

$$= \beta_{2(t+1)}^1 \alpha_{1(t+1)}^1 + \beta_{4(t+1)} \beta_{2t}^1 \alpha_{1t}^1$$

At time t+2,

$$M_{i(t+2)}^1 = \alpha_{0(t+2)}^1 + \ \alpha_{1(t+2)}^1 X_{i(t+2)} + e_{t+2}^{M1}$$

$$Y_{i(t+2)} = \beta_{0(t+2)} + \ \beta_{1(t+2)} X_{i(t+2)} + \textstyle\sum_{k=1}^{d} \beta_{2(t+2)}^k M_{i(t+2)}^k + \beta_{3(t+2)} X_{i(t+1)} + \beta_{4(t+2)} Y_{i(t+1)} +$$
$$\beta_{5(t+2)} X_{it} + \beta_{6(t+2)} Y_{it} + e_{t+2}^y$$

Assuming $e_{t+2}^{M1} \perp e_{t+2}^{y}$, $e_{t+1}^{y} \perp e_{t+2}^{y}$, $e_{t}^{y} \perp e_{t+2}^{y}$, we can prove:

$$E\left(\widehat{\beta_{2(t+2)}^{1}}\widehat{\alpha_{1(t+2)}^{1}} + \widehat{\beta_{4(t+2)}^{1}}\ \widehat{\beta_{2(t+1)}^{1}}\widehat{\alpha_{1(t+1)}^{1}} + (\widehat{\beta_{4(t+2)}^{1}}\widehat{\beta_{4(t+1)}^{1}} + \widehat{\beta_{6(t+2)}^{1}})\widehat{\beta_{2t}^{1}}\widehat{\alpha_{1t}^{1}}\right)$$

$$= E\left(\widehat{\beta_{2(t+2)}^{1}}\widehat{\alpha_{1(t+2)}^{1}}\right) + E\left(\widehat{\beta_{4(t+2)}^{1}}\ \widehat{\beta_{2(t+1)}^{1}}\widehat{\alpha_{1(t+1)}^{1}}\right) + E\left(\widehat{\beta_{4(t+2)}^{1}}\widehat{\beta_{4(t+1)}^{1}}\widehat{\beta_{2t}^{1}}\widehat{\alpha_{1t}^{1}}\right) +$$

$$E\left(\widehat{\beta_{6(t+2)}^{1}}\widehat{\beta_{2t}^{1}}\widehat{\alpha_{1t}^{1}}\right)$$

$$= \beta_{2(t+2)}^{1}\alpha_{1(t+2)}^{1} + \beta_{4(t+2)}^{1}\beta_{2(t+1)}^{1}\alpha_{1(t+1)}^{1} + (\beta_{4(t+2)}^{1}\beta_{4(t+1)}^{1} + \beta_{6(t+2)}^{1})\beta_{2}^{t}\alpha_{1}^{t}$$

For multiple mediators, we can prove unbiases by adding them together. For estimator of direct effect, we can prove its unbiases in the same method.

### S3. Parameters for each simulation scenario

Exposure $X_T$ followed a multinormal distribution, $(X_t,\ X_{t+1},\ X_{t+2})^T \sim MNV(\boldsymbol{\mu}, \Sigma)$, $\boldsymbol{\mu} = \begin{bmatrix} 0 \\ 0 \\ 0 \end{bmatrix}$, variance with moderate correlation $\Sigma = \begin{bmatrix} 1 & 0.6 & 0.5 \\ 0.6 & 1 & 0.6 \\ 0.5 & 0.6 & 1 \end{bmatrix}$ or high correlation $\Sigma = \begin{bmatrix} 1 & 0.9 & 0.8 \\ 0.9 & 1 & 0.9 \\ 0.8 & 0.6 & 1 \end{bmatrix}$. The mediator model at each time point was $M_{iT}^{1} = 1 + 0.4 \times X_{it} + \ \varepsilon_{T}^{1}$, where $\varepsilon_{T}^{1} \sim N(0,\ 1)$. The outcome model with a high proportion of variance explained by exposures, the mediator, and previous outcomes was specified as follows:

$Y_{it} = 1 + 0.7 \times X_{it} + 0.6 \times M_{it} + \ \varepsilon_{t}^{Y}$, where $\varepsilon_{t}^{Y} \sim N(0,\ 1)$

$Y_{i(t+1)} = 1 + 0.4 \times X_{i(t+1)} + 0.5 \times M_{i(t+1)} - 0.2 \times X_{it} + 0.5 \times Y_{it} +$
$\varepsilon_{t+1}^{Y}$, where $\varepsilon_{t+1}^{Y} \sim N(0,\ 1)$

$Y_{i(t+2)} = 1 + 0.4 \times X_{i(t+2)} + 0.5 \times M_{i(t+2)} - 0.2 \times X_{i(t+1)} + 0.5 \times Y_{i(t+1)} - 0.1 \times X_{it} +$

$0.2 \times Y_{it} + \varepsilon^Y_{t+2}$, where $\varepsilon^Y_{t+2} \sim N(0,\ 1)$

The outcome model with low variance explained was as follows:

$Y_{it} = 1 + 0.3 \times X_{it} + 0.2 \times M_{it} + \varepsilon^Y_t$, where $\varepsilon^Y_t \sim N(0,\ 1)$

$Y_{i(t+1)} = 1 + 0.3 \times X_{i(t+1)} + 0.2 \times M_{i(t+1)} - 0.1 \times X_{it} + 0.1 \times Y_{it} +$

$\varepsilon^Y_{t+1}$, where $\varepsilon^Y_{t+1} \sim N(0,\ 1)$

$Y_{i(t+2)} = 1 + 0.2 \times X_{i(t+2)} + 0.2 \times M_{i(t+2)} - 0.1 \times X_{i(t+1)} + 0.2 \times Y_{i(t+1)} - 0.05 \times X_{it} +$

$0.1 \times Y_{it} + \varepsilon^Y_{t+2}$, where $\varepsilon^Y_{t+2} \sim N(0,\ 1)$

### S4. Comparison to cross-lagged panel model

Cross-lagged panel model is one of the common structural equation models for longitudinal mediation analysis. Cole and Maxwell proposed relationship for exposure X, mediator M and outcome Y as follows[1]:

$X_{t+1} = \beta_{X,t} X_t + \varsigma_{X,t}$

$M_{t+1} = \beta_{M,t} M_t + \beta_{X,t} X_t + \varsigma_{M,t}$

$Y_{t+2} = \beta_{Y,t+1} Y_{t+1} + \beta_{M,t+1} M_{t+1} + \beta_{X,t} X_t + \varsigma_{Y,t}$

Where t, t+1 and t+2 are three time points, and $\varsigma_{X,t}$ $\varsigma_{M,t}$ and $\varsigma_{Y,t}$ are error terms of the three models. The indirect effect is $\beta_{X,t}\beta_{M,t+1}$. The cross-lagged panel model improves upon cross-sectional analyses by incorporating time lags. Compared to this model, our proposed model adjusts for $M_{t+2}$ instead of $M_{t+1}$, adjust for $X_{t+2}$ instead of $X_t$, and additionally includes $Y_t$ as a covariate in the model for $Y_{t+2}$. This indirect effect is constructed from X-M relationship from time t and M-Y relationship at time t+1, while our cumulative indirect effect includes indirect effect from all the time points. We used simulation scenario 1 dataset to compare the two methods. The simulation showed that cross-lagged panel model was unable to accurately capture the true cumulative indirect effect at time t+2, estimating an effect of -0.01 compared to the true value of 0.41.

**S5. Unmeasured confounding**

Causal mediation analysis was developed by Robins and Greenland[2] and Pearl[3]. For identification of natural direct and indirect effect, VanderWeele and Vansterrlandt[4] proposed unmeasured confounding assumption: i) no measured confounding for exposure and outcome (X-Y); ii) no measured confounding for mediator and outcome (M-Y); ii) no measured confounding for exposure and mediator (X-M); iv) no mediator-outcome confounder that is itself affected by the exposure. For future extension of our cumulative model to causal mediation analysis framework, we assess how first three confounder affects the estimation of cumulative indirect effect. For simplicity, we consider a single mediator when analyzing the bias caused by unmeasured confounders (**Supplementary Figure 1**). The true models with unmeasured confounders are as follows:

$$M_{it}^{1} = \alpha_{0t}^{1} + \alpha_{1t}^{1} X_{it} + K_{XM} C_{XM} + e_{t}^{M1}$$

$$M_{i(t+1)}^{1} = \alpha_{0(t+1)}^{1} + \alpha_{1(t+1)}^{1} X_{i(t+1)} + K_{XM} C_{XM} + e_{(t+1)}^{M1}$$

$$M_{i(t+2)}^{1} = \alpha_{0(t+2)}^{1} + \alpha_{1(t+2)}^{1} X_{i(t+2)} + K_{XM} C_{XM} + e_{(t+2)}^{M1}$$

$$Y_{it} = \beta_{0t} + \ \beta_{1t}X_{it} + \beta_{2t}^{1}M_{it}^{1} + K_{XY}C_{XY} + K_{MY}C_{MY} + e_{t}^{y}$$

$$Y_{i(t+1)} = \beta_{0(t+1)} + \ \beta_{1(t+1)}X_{i(t+1)} + \beta_{2(t+1)}^{1}M_{i(t+1)}^{1} + \beta_{3(t+1)}X_{it} + \beta_{4(t+1)}Y_{it} + K_{XY}C_{XY}$$

$$+K_{MY}C_{MY} + e_{t+1}^{y}$$

$$Y_{i(t+2)} = \beta_{0(t+2)} + \ \beta_{1(t+2)}X_{i(t+2)} + \beta_{2(t+2)}^{1}M_{i(t+2)}^{1} + \beta_{3(t+2)}X_{i(t+1)} + \beta_{4(t+2)}Y_{i(t+1)}$$

$$+\beta_{5(t+2)}X_{it} + \beta_{6(t+2)}Y_{it} + K_{XY}C_{XY} + K_{MY}C_{MY} + e_{t+2}^{y}$$

Without adjusting for unmeasured confounders, the observed models have the bias for each parameter:

$$M_{it}^{1} = \alpha_{0t}^{1} + \left(\alpha_{1t}^{1} + \delta_{XM\alpha_{1t}}\right)X_{it} + \varepsilon_{t}^{M1}$$

$$M_{i(t+1)}^{1} = \alpha_{0(t+1)}^{1} + \left(\alpha_{1(t+1)}^{1} + \delta_{XM\alpha_{1(t+1)}}\right)X_{i(t+1)} + \varepsilon_{(t+1)}^{M1}$$

$$M_{i(t+2)}^{1} = \alpha_{0(t+2)}^{1} + \left(\alpha_{1(t+2)}^{1} + \delta_{XM\alpha_{1(t+2)}}\right)X_{i(t+2)} + \varepsilon_{(t+2)}^{M1}$$

$$Y_{it} = \beta_{0t} + \left(\ \beta_{1t} + \delta_{XY\beta_{1t}}\right)X_{it} + \left(\beta_{2t}^{1} + \delta_{MY\beta_{2t}}\right)M_{it}^{1} + \varepsilon_{t}^{Y}$$

$$Y_{i(t+1)} = \beta_{0(t+1)} + \ (\beta_{1(t+1)} + \delta_{XY\beta_{1(t+1)}})X_{i(t+1)} + (\beta_{2(t+1)}^{1} + \delta_{MY\beta_{2(t+1)}})M_{i(t+1)}^{1} + (\beta_{3(t+1)}$$

$$+\delta_{XY\beta_{3(t+1)}})X_{it} + (\beta_{4(t+1)} + \delta_{XY\beta_{4(t+1)}} + \delta_{MY\beta_{4(t+1)}})Y_{it} + \varepsilon_{(t+1)}^{Y}$$

$$Y_{i(t+2)} = \beta_{0(t+2)} + \ (\beta_{1(t+2)} + \delta_{XY\beta_{1(t+2)}})X_{i(t+2)} + (\beta_{2(t+2)}^{1} + \delta_{MY\beta_{2(t+2)}})M_{i(t+2)}^{1} + (\beta_{3(t+2)}$$

$$+\delta_{XY\beta_{3(t+2)}})X_{i(t+1)} + (\beta_{4(t+2)} + \delta_{XY\beta_{4(t+2)}} + \delta_{MY\beta_{4(t+2)}})Y_{i(t+1)}$$

$$+(\beta_{5(t+2)} + \delta_{XY\beta_{5(t+2)}})X_{it} + (\beta_{6(t+2)} + \delta_{XY\beta_{6(t+2)}} + \delta_{MY\beta_{6(t+2)}})Y_{it} + \varepsilon_{(t+2)}^{Y}$$

where $\delta_{XM\alpha_{1t}}$, $\delta_{XM\alpha_{1(t+1)}}$, $\delta_{XM\alpha_{1(t+2)}}$ are bias in $\alpha_{1t}^1$ , $\alpha_{1(t+1)}^1$, $\alpha_{1(t+2)}^1$ , respectively, due to the unmeasured X-M confounder, $\delta_{MY\beta_{2t}}$, $\delta_{MY\beta_{2(t+1)}}$, $\delta_{MY\beta_{2(t+2)}}$are bias in $\beta_{2t}^1$, $\beta_{2(t+1)}^1$, $\beta_{2(t+2)}^1$ due to the unmeasured M-Y confounder, similarly $\delta_{MY\beta_4}$ and $\delta_{MY\beta_6}$ at each time point are bias due to unmeasured M-Y confounder, and $\delta_{XY\beta_1}$ , $\delta_{XY\beta_3}$, $\delta_{XY\beta_5}$ are bias due to the unmeasured X-Y confounder. Therefore, the cumulative direct and indirect effects have the corresponding biases.

Specifically, we simulated the data from the model with an exposure-mediator confounder as shown in Supplementary Figure 1. The $[C_t, C_{t+1}, C_{t+2}]$ followed a multinormal distribution, at each time point the mean of exposure was zero and variance was one. The auto-regressive Pearson correlation between exposure $[X_t, X_{t+1}, X_{t+2}]$ and unmeasured $[C_t, C_{t+1}, C_{t+2}]$ was moderate correlated (**Supplementary Table 2**). The true mediator model with X-M confounder at time point t is $M_{it} = 1 + 0.4 \times X_{it} + K_t C_{it} + \varepsilon_t^1, \ \varepsilon_t^1 \sim N(0,\ 1)$, and similar model for time points t+1 and t+2. The strength of $K_t$ represented the magnitude of bias in $\alpha_{1t}$ resulted from unmeasured confounder. The magnitude of confounding was quantified by $\frac{\delta_{\alpha_{1t}}}{\alpha_{1t}}$. We assumed there were no unmeasured confounders for the X-Y, or Y-M relationships. We performed 1,000 independent simulation replicates to evaluate the bias on exposure under different confounding strength.

Supplementary Figure 2 illustrated that the existence of unmeasured confounding triggered in bias, ranging from 5% to 100%, in the estimated cumulative indirect effect across time points. Furthermore, RMSE increased progressively from 0.02 to 0.4 with the strength of the unmeasured confounding increasing, indicating a notable decline in estimation precision. Coverage probability dropped from 95% to 0% as confounding strengthened, with coverage hitting zero when estimated $\alpha_{1t}$was 50% of true $\alpha_{1t}$. The results indicated that our proposed cumulative indirect effect estimator was not robust to the effect of an unmeasured confounders between exposure and mediator (**Supplementary Figure 3**).

**Supplementary Table 1. Selected CpG and its coefficient from elastic net regression**

| CpG | UCSC.Gene | Chromosome | Position | Coefficient |
|---|---|---|---|---|
| cg00716257 | JDP2 | 14 | 75897417 | -4.35 |
| cg01687189 | GNPTAB | 12 | 102225365 | -3.41 |
| cg03359362 | SLC1A5 | 19 | 47289611 | -13.15 |
| cg03575969 | STARD10 | 11 | 72492171 | -2.70 |
| cg05130679 | AMOTL1 | 11 | 94502824 | -4.53 |
| cg05991220 | ITIH1 | 3 | 52813859 | 1.96 |
| cg06690548 | SLC7A11 | 4 | 139162808 | -9.18 |
| cg15804598 | HEXIM1 | 17 | 43224418 | -5.71 |
| cg23534245 | BMP2 | 20 | 6751435 | -3.44 |
| cg24848615 | NFIC | 19 | 3368396 | -10.21 |
| cg25518868 | DIAPH1 | 5 | 140984057 | -0.87 |

**Supplementary Table 2. Pearson correlation between exposures and unmeasured confounder**

| | $X_t$ | $X_{t+1}$ | $X_{t+2}$ | $C_t$ | $C_{t+1}$ | $C_{t+2}$ |
|---|---|---|---|---|---|---|
| $X_t$ | 1 | | | | | |
| $X_{t+1}$ | 0.6 | 1 | | | | |
| $X_{t+2}$ | 0.5 | 0.6 | 1 | | | |
| $C_t$ | 0.5 | 0.3 | 0.1 | 1 | | |
| $C_{t+1}$ | 0.3 | 0.5 | 0.3 | 0.6 | 1 | |
| $C_{t+2}$ | 0.1 | 0.3 | 0.5 | 0.5 | 0.6 | 1 |

**Supplementary Figure 1. Unmeasured confounders in the cumulative mediation analysis**
Examples for unmeasured confounders between exposure and mediator (A), mediator and outcome (B), exposure and outcome (C) at each time point. Unmeasured confounders introduce bias in estimating association between exposure, mediator and outcome, which in turn lead to biased estimation of cumulative indirect effect.

A. **An example for unmeasured confounder between exposure and mediator**

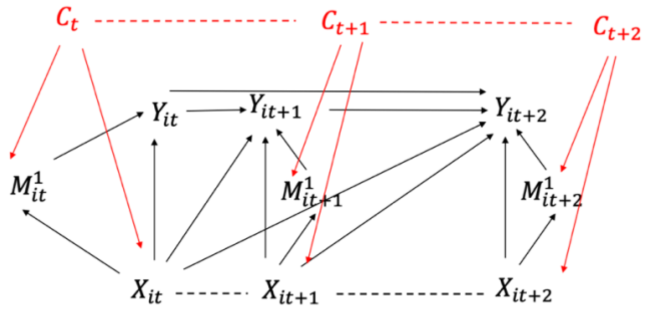


B. **An example for unmeasured confounder between mediator and outcome**

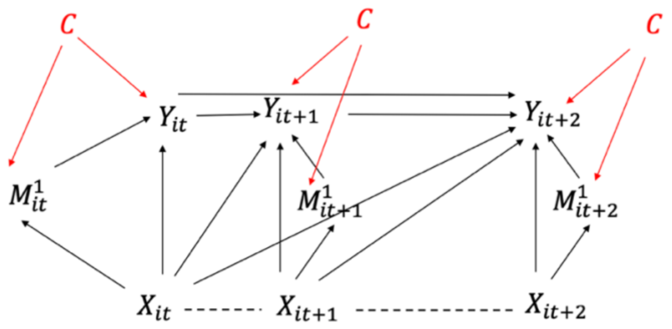


C. **An example for unmeasured confounder between exposure and outcome**

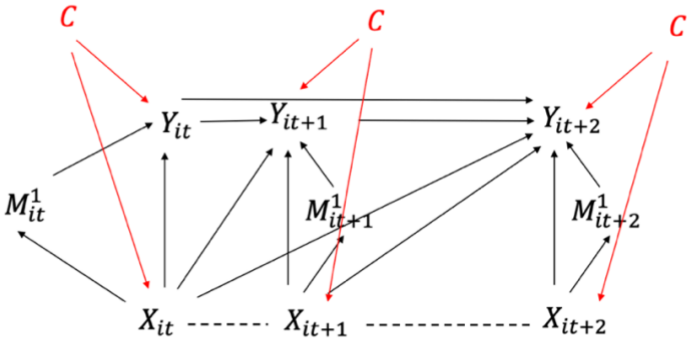

**Supplementary Figure 2. Bias for cumulative indirect effect under unmeasured confounder between exposure and mediator**

The auto-regressive Pearson correlation between exposure $X_T$ and unmeasured $C_T$ was moderate correlated. The true mediator model with X-M confounder at each time point is $M_{iT} = 1 + 0.4 \times X_{iT} + K_T C_{iT} + \varepsilon_T^1, \ \varepsilon_T^1 \sim N(0,\ 1)$. The strength of $K_T$ represented the magnitude of bias in $\alpha_1^T$ (i.e., $\delta_{\alpha_1}^t = \widehat{\alpha_1^t} - \alpha_1^t$) resulted from unmeasured confounder. The magnitude of confounding was quantified by $\frac{\delta_{\alpha_1}^t}{\alpha_1^t}$. We performed 1,000 independent simulation replicates to evaluate the bias on exposure under different confounding strength.

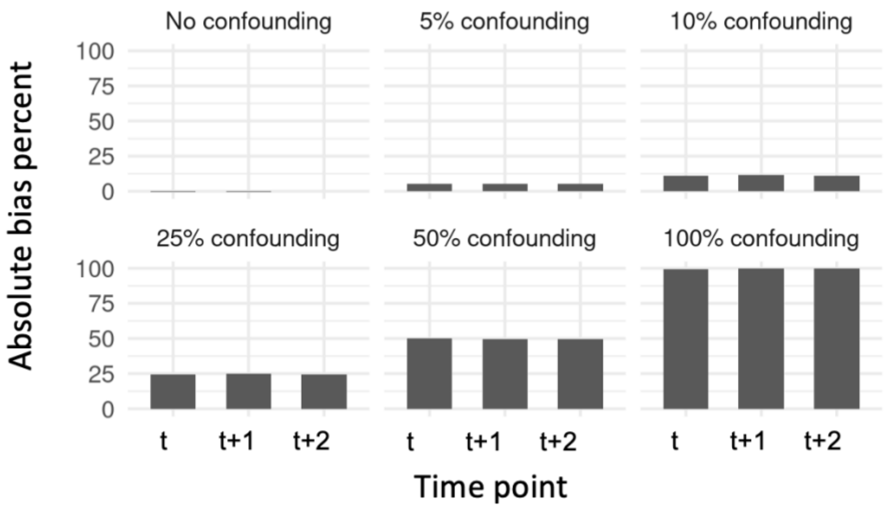

**Supplementary Figure 3. Coverage probability for cumulative indirect effect 95% confidence interval under unmeasured confounder between exposure and mediator**

Standard error of cumulative indirect effect was calculated using three methods (i.e., bootstrap procedure, delta method and exact variance). At the time t+2, the exact variance calculation was too complex and therefore omitted from the figure. The 95% confidence interval of cumulative indirect effect was constructed as estimated cumulative indirect effect $\pm$ 1.96 $\times$ standard error.

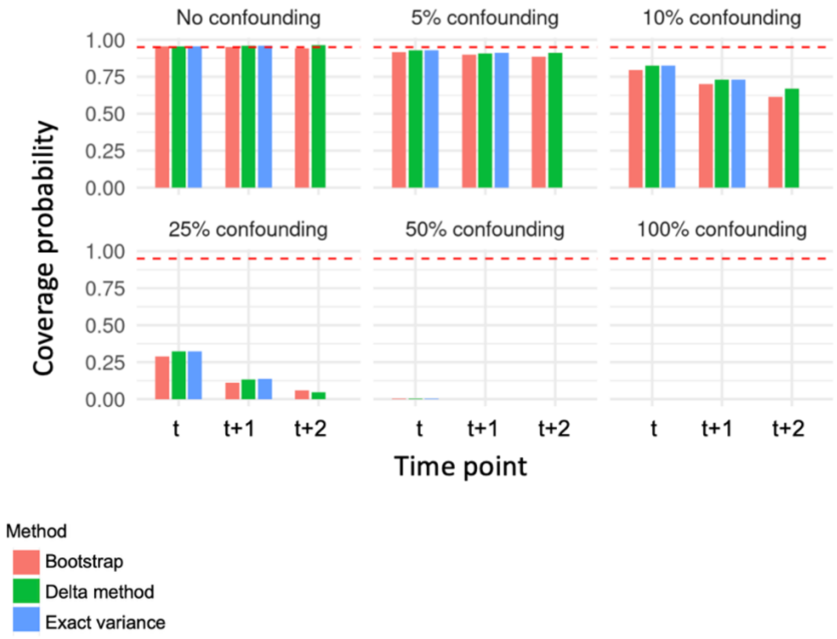
